# Novel Mutations in Human Luteinizing Hormone Beta Subunit Related to Polycystic Ovary Syndrome among Sudanese Women

**DOI:** 10.1101/2020.10.11.20208926

**Authors:** Nidal Essa, Suliman Osman, Salah Jemaah, Mohamed A. Hassan, Rashid Eltayeb

## Abstract

**Introduction:** Polycystic ovary syndrome (PCOS) is a common disorder that is not fully understood. Multiple hormonal and metabolic factors impact on disease pathophysiology resulting in various phenotypic characteristics among the PCOS population. Luteinizing hormone beta subunit (LHB) (protein ID P01229) is mapped on (chr19p13.3) and consists of three exons. Luteinizing hormone (LH) has a central role in stimulation ovarian steroidogenesis, in particular androgen production, and the promotion of ovulation.

**Objectives:** To determine if genetic variations of LHB are associated with PCOS among Sudanese families.

**Methods:** A prospective laboratory based cross-sectional study to examine genetic mutations in LHB that associate with PCOS in families (cases; n=35 families, 90 females and controls; n=11 families, 30 females) in Khartoum State, Sudan. Quantitative enzyme linked immuno-sorbent assay (ELISA) and polymerase chain reaction (PCR) with Sanger sequencing were used to analyze biochemical parameters and detect polymorphisms. Protein structure and function bioinformatics analysis was conducted using standard software.

**Results:** PCOS cases had significantly different biochemical parameters from the controls (LH: p<0.001; testosterone: p<0.001; fasting glucose: p=0.02; insulin: p=0.01; triglycerides: p=0.03; total cholesterol: p<0.001; high density lipoprotein (HDL): p=0.012;low density lipoprotein (LDL): p<0.001). There were no differences in follicle stimulating hormone (FSH) (p=0.984) or prolactin (p=0.068). Sanger sequencing revealed 5 single nucleotide polymorphisms (rs5030775, A18T; rs746167425, R22K; rs1800447, W28R; rs35270001, H30R; and rs34349826, I35T) located on (exon 2) of *LHB* gene that were statistically correlated with serum LH, Testosterone and insulin levels among PCOS families.

**Conclusion:** This is the first molecular family-based study in Sudan exploring the genetics of the *LHB* gene in women manifesting PCOS. These novel mutations give further information about the role of genetic inheritance and may explain some of the altered ovarian function and responses in women with PCOS.

## Introduction

Polycystic ovary syndrome (PCOS) is a disorder which is associated with a hormone imbalance, anovulation and multiple paused follicles in the ovaries that is linked to disparate metabolic disorders.^**1**,**2**^ It has a complex etiology with genetic, epigenetic and environmental factors. Adherence to the three common diagnostic criteria of increased androgens, polycystic ovaries on ultrasound scan and anovulation (1990 National Institutes of Health’s (NIH), the Rotterdam criteria and the Androgen Excess Society), which cover the different phenotypic features of PCOS, suggests a prevalence of 1.6-18% among women of reproductive age.^2-4^

There are epigenetic factors that can promote PCOS. In various animal models, and in women, increased exposure of the developing fetus to increased androgen concentrations before birth can result in developmental programming of the features of PCOS. Prenatal androgens result in dynamic changes in gene expression, resulting hyperinsulinemia, adipocyte dysfunction, visceral obesity, and PCOS in offspring.^**9**, **10**^ In addition, there are environmental factors involved in PCOS. An increase of weight can unmask a PCOS phenotype in adulthood.

However, there are genetic elements of PCOS, although the genes involved remain poorly characterized. Various familial aggregation studies suggest that PCOS is an autosomal dominant syndrome and there is an association in both sisters and brothers of women with PCOS with infertility problems. In addition monozygotic and dizygotic twin studies suggest a genetic contribution to PCOS. Recent genome-wide association studies (GWAS) in Han Chinese women with PCOS identified 11 genetic loci that related to PCOS. These loci were in genes involved in steroidogenesis, steroid hormone action, gonadotrophin action, regulation of internal secretion, energy balance, chronic inflammation, and other pathways relevant to PCOS.^**11-16**^

PCOS is associated with an exaggerated LH pulse frequency and amplitude that enhances androgen secretion. In addition, there are alterations in the androgen synthetic pathway that associate with enhanced ovarian and adrenal androgen production^**17-19**^. Moreover, excessive androgen secretion results on many types of hormonal and metabolic abnormalities, which are characterized by hirsutism, anovulation, clinical hyperandrogenism, hyperinsulinemia, insulin resistance (IR), and obesity, which are all more common in PCOS^**1-3**^. Therefore, assessment of hormonal profile among PCOS population using high-quality techniques such as liquid chromatography tandem mass spectrometry (LC-MS/MS), plays a key diagnostic marker of PCOS.

As androgens are driven by LH, we focused on the *LHB* gene. We aimed to examine the relationship between single nucleotide polymorphisms (SNPs) of the *LHB* gene with the clinical features of PCOS among Sudanese women within the reproductive age.

## Methodology

This study was approved by the national ethics committee - Federal Ministry of Health (3-12-19). The inclusion criteria for PCOS families were women diagnosed with PCOS, according to the Rotterdam criteria that had one or two sisters diagnosed as PCOS within the reproductive age (20-40) years. In control families neither the woman nor any sisters had any features of PCOS. Any women below the reproductive age or with family history of adrenal hyperplasia, hyperprolactinemia, Cushing syndrome, thyroid disorders, ovarian cancer or breast cancer and causes of infertility other than PCOS were excluded from the study.

This prospective laboratory based cross-sectional study involved blood sampling carried out in different fertility centers as well as different localities (families’ homes) in Khartoum State. Hormonal profile estimations were carried out using validated tests within the clinical service laboratory with quantitative enzyme linked immunosorbent assay (ELISA) and enzymatic methods for measurement of fasting blood glucose and lipid profile. DNA extraction from whole blood was carried out using a commercial kit according to the manufacturer’s instructions. Primer 3 was used to design the *LHB* primers (Table 1). Routine polymerase chain reaction (PCR) and gel electrophoresis was used for band amplification and visualization. The Sanger dioxnucleotidue sequencing method was used for genotyping and the bioinformatics software (BLAST, USCF Chimera, Project Hope and Raptor X) was used for protein structure, function and prediction.

**Table 1:**
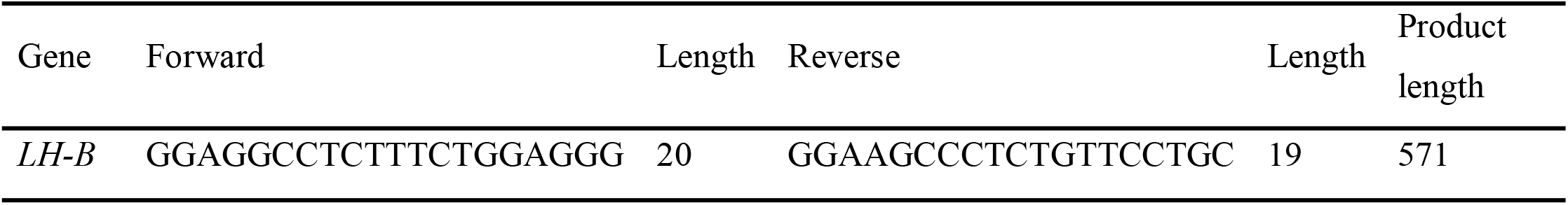
The *LHB* gene primers for PCR protocol.

Descriptive and biological information about study groups were analyzed by SPSS version 23 using descriptive analysis for age, BMI, and biochemical analysis. T-tests were used for statistical significance after testing for normality, Chi square test were used for proportion analysis and Person correlation were used for statistical association. Statistical significance was p<0.05.

## Results

The patient characteristics and biochemical parameters are shown in (Table 2). There was no difference in age and BMI, serum levels of FSH and PRL assessment between the groups. However, there were clear increases in LH and testosterone concentrations (p<0.001) in the PCOS group. This meant that the LH: FSH ratio was significantly elevated in women with PCOS (P<0.001). In addition, there was evidence of insulin resistance and dyslipidaemia in the women with PCOS (Table 2). Women with PCOS suffered from signs of hyperandrogenism with hirsutism (42.9%), acne (54.5%) and seborrhea (30%) (Table 3) and testosterone concentrations were significantly linked to menstrual irregularity (Table 4). This highlights the validity of the cohort of PCOS patients when compared to controls.

**Table (2).** Characteristics of PCOS and controls cohorts:

| **Parameters** | **Study groups** | **Mean± SD** | ***p-value*** |
| --- | --- | --- | --- |
| **Age. yrs.** | Case | 35.3±13,9 | 0.283 |
|  | Control | 37±18,1 |  |
| **BMI** | Case | 28.6±112,9 | 0.529 |
|  | Control | 26.6±59,3 |  |
| **FBG/mmol/l** | Case(n=90) | 6.2 ±1.8 | 0.020* |
|  | Control(n=30) | 5.4±0.9 |  |
| **LH (ng/dl)** | Case (n=90) | 17.7±12.3 | 0.000* |
|  | Control(n=30) | 9.7±5.4 |  |
| **FSH (ng/dl)** | Case(n=90) | 5.4±4.6 | 0.984 |
|  | Control(n=30) | 5.4±2.9 |  |
| **PRL (ng/dl)** | Case(n=90) | 12.4±8.2 | 0.068 |
|  | Control(n=30) | 8.0±6.2 |  |
| **LH/FSH ratio** | Case(n=90) | 5.5±6.1 | 0.000* |
|  | Control(n=30) | 1.9±0.8 |  |
| **TG (mmol/l)** | Case(n=90) | 1.5±0.6 | 0.030* |
|  | Control(n=30) | 1.1±0.3 |  |
| **T.Chol (mmol/l)** | Case(n=90) | 4.1±1.5 | 0.000* |
|  | Control(n=30) | 2.2±0.4 |  |
| **HDL(mmol/l)** | Case(n=90) | 1.2±0.2 | 0.012* |
|  | Control(n=30) | 1.4±0.2 |  |
| **LDL(mmol/l)** | Case(n=90) | 2.3±1. 6 | 0.000* |
|  | Control(n=30) | 0.27±0.5 |  |
| **Insulin (ng/dl)** | Case(n=90) | 15.2±8.4 | 0.010* |
|  | Control(n=30) | 7.7±6.2 |  |
| **Testosterone (ng/dl)** | Case(n=90) | 33.6±18.0 | 0.000* |
|  | Control(n=30) | 10.5±7.0 |  |

\* The mean difference is significant at the 0.050 level

**Table (3):** The association between clinical hyperandrogenism and menstrual cycle status among study groups.

| Clinical Hyperandrogenism |  | Status of MC |  | *P-value* |
| --- | --- | --- | --- | --- |
|  |  | Regular | Irregular |  |
| **Hirsutism** | Yes | 15% | 24% | 0.000* |
|  | No | 42% | 17% |  |
| **Acne** | Yes | 25% | 27% | 0.040* |
|  | No | 32.2% | 15% |  |
| **Alopecia** | Yes | 17% | 18 | 0.040* |
|  | No | 42.2% | 22.2% |  |
| **Seborrhea** | Yes | 17% | 17.7% | 0.130 |
|  | No | 42.2% | 24.4% |  |

\* The mean difference is significant at the 0.05 level.

**Table (4):** The association between hormonal profile and menstrual cycle status among study groups.

| Parameters | Status of MC |  | *P-value* |
| --- | --- | --- | --- |
| | Regular<br>($\bar{x} \pm SD$) | Irregular<br>($\bar{x} \pm SD$) | |
| **LH (ng/dl)** | 16.9±127,9 | 16,079±106,9 | 0.741 |
| **FSH (ng/dl)** | 5,077±41,2 | 5,750±47,4 | 0.476 |
| **PRL (ng/dl)** | 11,708±75,4 | 11.932±88,8 | 0.900 |
| **Insulin(ng/dl)** | 12.90±8,3 | 15.84±8,6 | 0.108 |
| **Testosterone (ng/dl)** | 26.00±17,8 | 36.18±18,5 | 0.011* |

\* The mean difference is significant at the 0.05 level.

DNA was extracted and sequenced from the cases and controls and the chemical structure of the LH protein determined (Figure 1). Within the PCOS families there were five SNPs that were identified within exon (2) of LHβ (Figure 2, Table 5). These were significantly associated with serum levels of LH, testosterone and menstrual irregularity (Tables 6, 7).

**Table (5).**
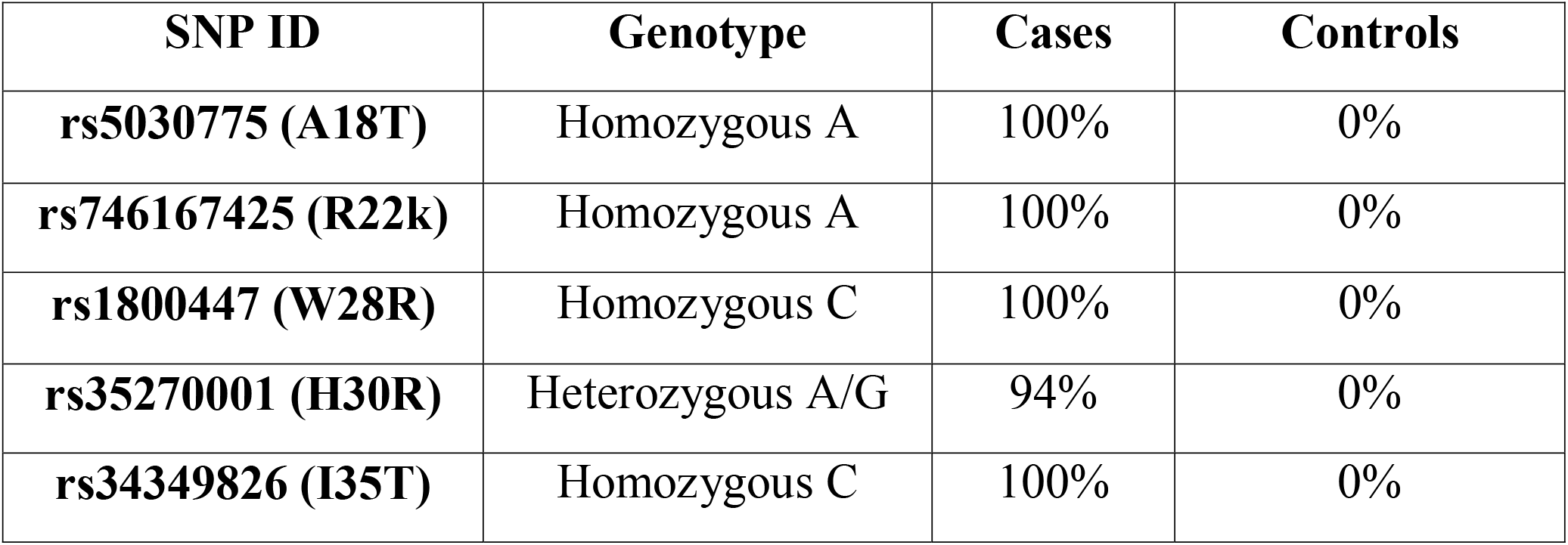
Distribution of LHβ genotypes and allele frequencies in healthy controls and patients with PCOS

| SNP ID | Genotype | Cases | Controls |
| --- | --- | --- | --- |
| rs5030775 (A18T) | Homozygous A | 100% | 0% |
| rs746167425 (R22k) | Homozygous A | 100% | 0% |
| rs1800447 (W28R) | Homozygous C | 100% | 0% |
| rs35270001 (H30R) | Heterozygous A/G | 94% | 0% |
| rs34349826 (I35T) | Homozygous C | 100% | 0% |

**Table (6):**
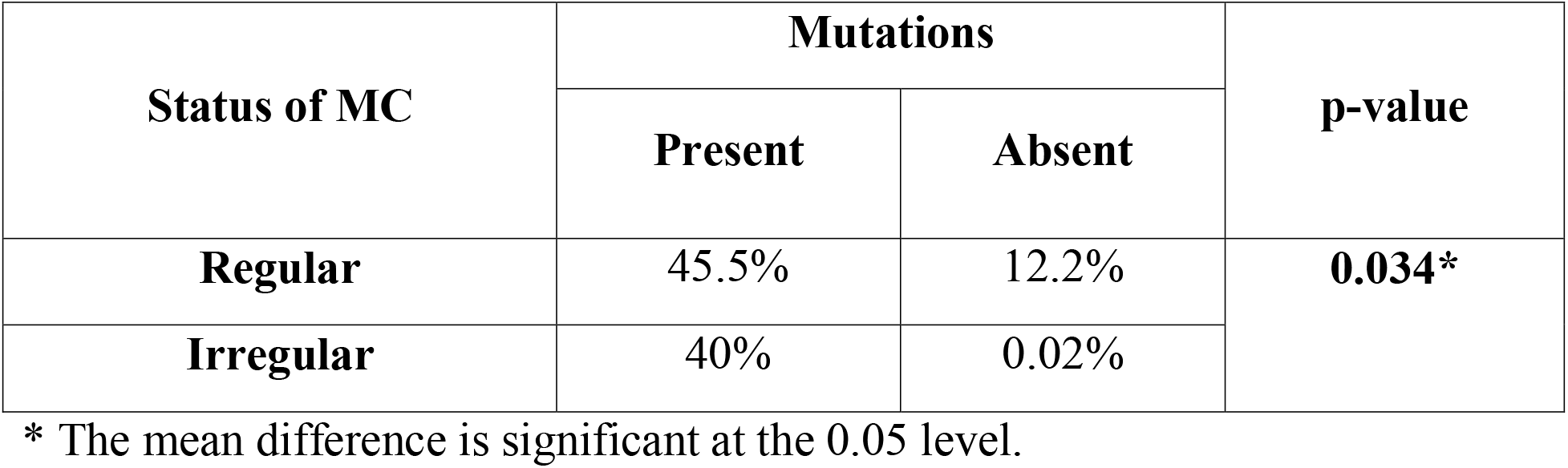
The association of LHβ polymorphisms and menstrual irregularity among study groups:

**Table (7):** The association of LHβ polymorphisms and hormonal profile among study groups:

| Parameters | Mutation |  | 95% CI |  | p-value |
| --- | --- | --- | --- | --- | --- |
|  | Present | Absent |  |  |  |
| **LH (ng/dl)** | 17.732±23.2 | 9.6±53.5 | 11.395 | 149.870 | .023* |
| **FSH (ng/dl)** | 5.3±46.11 | 5.3±28.61 | -26.038 | 26.567 | .984 |
| **Prolactin (ng/dl)** | 12.442±82.2 | 8.0±61.8 | -.3297 | 91.821 | .068 |
| **Insulin (ng/dl)** | 15.3±8.4 | 7.6±6.1 | 2.673 | 12.410 | .003* |
| **Testosterone (ng/dl)** | 33.6±18.0 | 10.7±7.0 | 17.369 | 28.827 | .000* |

**Figure (1):**
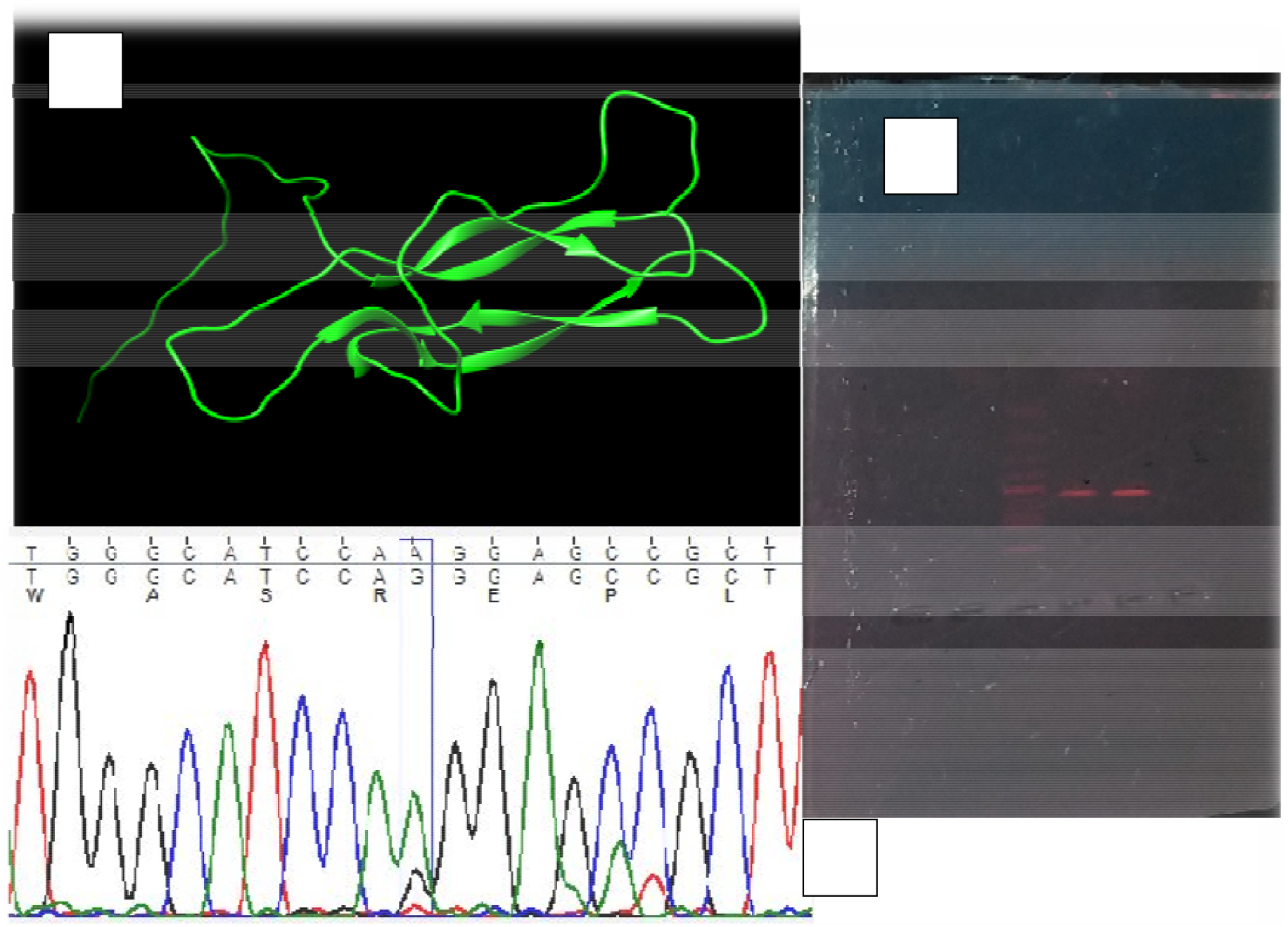
(A)The normal chemical structure of LH protein, (B) The chromatogram graph for the sequenced protein, and (C) The gel electrophoresis showing the LH band.

**Figure (2).**
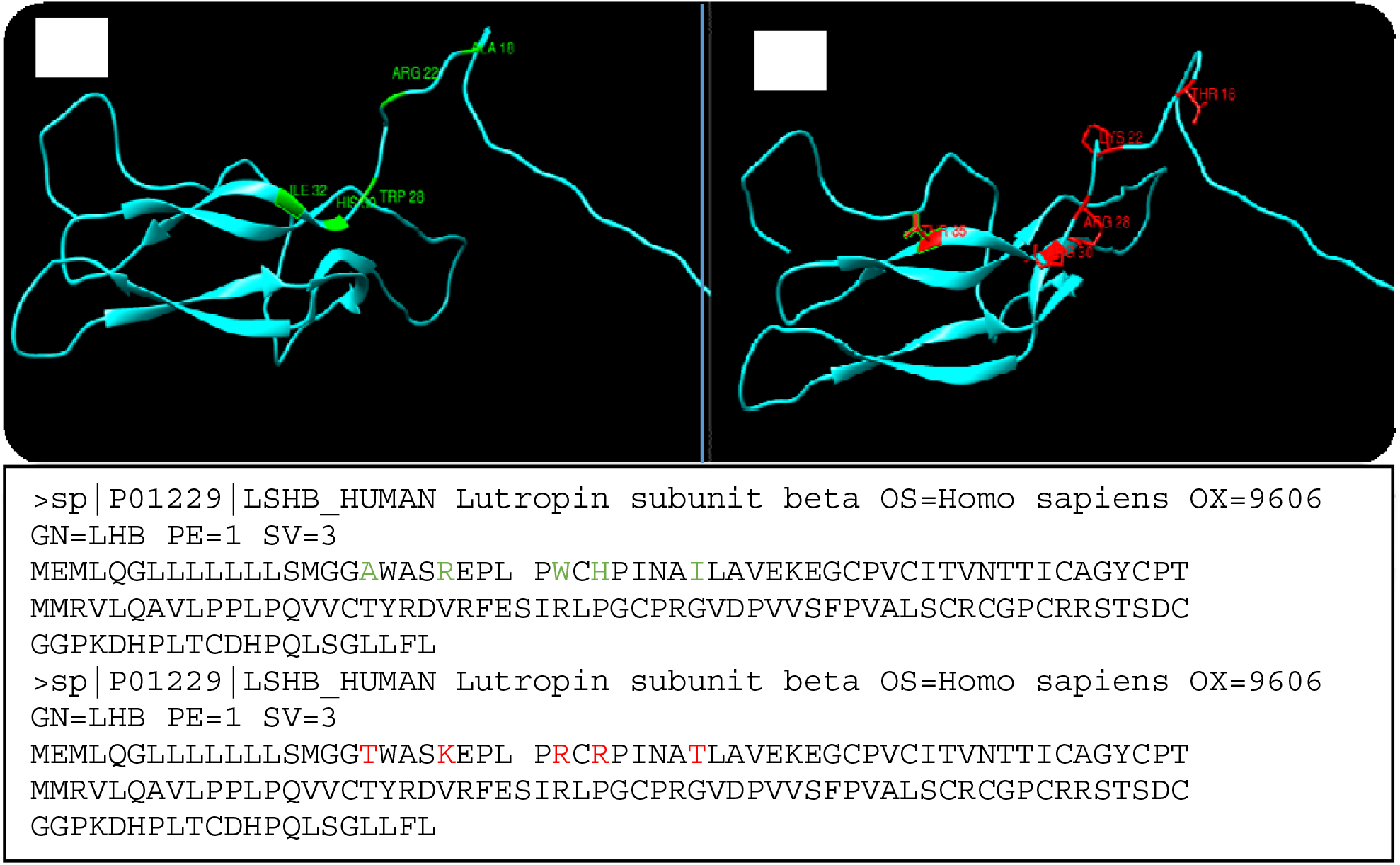
Example of normal (A) and mutated amino acids (red color) on sequenced exon (2) LHβ gene (B).

## 4. Discussion

In this study we investigated if different biochemical parameters were associated with polymorphisms of the *LHB* gene among Sudanese families who had more than one PCOS sufferer and compared this to control families without women with PCOS. The biochemical parameters within the PCOS group were consistent with PCOS and showed that they were different to the control group. We identified five polymorphisms in the LHB gene in the families with PCOS in Sudan. There were no polymorphisms in the control cases.

In this study the biochemical parameters were within the normal ranges but significantly different from the control group of the study, except BMI, serum levels of FSH and PRL. The cases and controls were clearly different symptomatically. Women with PCOS were more likely to suffer from menstrual irregularity (46.8%) and signs of clinical hyperandrogenism; facial hirsutism (42.9%), Acne (54.5%), Seborrhea (33.3%) and Alopecia (35.6%) (Esmaeilzadeh et al., 2014; Patel, 2018) than controls. They also showed different biochemical profiles with consensus to the international hallmarks of PCOS of hormonal imbalance (Act, 2018; Azziz, 2016; Escobar-Morreale, 2018; Harris et al., 2017).

In agreement with many previous studies the PCOS cohort also showed dyslipidemia (Esmaeilzadeh et al., 2015; Shoaib et al., 2015; Bennal et al., 2014). In contrast, Sadeghi et al., 2014 suggested that this was obesity dependent. They showed that there is no association between serum lipid profile, total cholesterol level, LDL cholesterol and HDL cholesterol (p-value =0.86, 021, 0.76 respectively) and PCOS except in the term of TG (p-value =0.0.4) in PCOS women with BMI > 25 kg/m^2^. While only obese PCOS women showed significant difference on serum TC as well as LDL-C (p=0.000) when compared with overweight/normal PCOS indicating that obesity, rather than PCOS per se, makes women more susceptible to dyslipidemia and cardiovascular disease (Castelo-Branco et al., ; Spałkowska et al., 2018). In our study there was no difference in BMI, and the mean BMI was only slightly overweight, but women with PCOS demonstrated dyslipidaemia.

In this cohort the higher glucose and insulin suggests the PCOS women are more predisposed to T2DM on their future life similar. This finding is similar to Forslund et al., (2020) and Rashidi (2018) who showed increased risk across different BMI groups. Obesity is a risk factor for impaired glucose tolerance as is PCOS with increased androgen concentrations. There is an adverse effects of obesity on serum testosterone level which is an inverse correlation among PCOS women with BMI ≥ 30 (p=0.014) while lean PCOS women had positive correlation (p=0.009). This suggests that the interaction of obesity and hyperandrogenism in the development of impaired glucose tolerance is complex (Spalkowska et al., 2018; Saxena et al.., 2013). Hyperandrogenemia is responsible for accumulation of the adipose tissue in the upper body (android fat distribution pattern; Good et al.). Interestingly Aydin et al., concluded that no differences in body composition between lean women with PCOS and healthy controls. Overall the influences of PCOS and obesity and their interaction on fat distribution and function requires further study.

We report that sisters of affected women have irregular menstrual cycles and this agrees with the Turkish study that showed that sisters had menstrual irregularity and larger ovaries with higher serum androstenedione and dehydroepiandrosterone sulfate levels than sisters without PCO, suggesting a spectrum of clinical phenotypes in PCO families. This parallels what was reported in a multicenter study with (1466) subjects, of whom (363) had PCOS and (79) polycystic ovaries (PCO) without other symptoms of PCOS. This is further evidence for PCOS clustering in families.

We examined the LHB gene as LH-stimulated androgen synthesis is fundamental to PCOS. We found that the LHB polymorphisms (rs1800447 (W28R) and rs34349826 (I35T) homozygotes (CC) had an association with menstrual irregularities with infertility, and elevation of serum testosterone level (Furui et al., 1994), whereas the Finnish subjects whom were homo- or heterozygous for the polymorphic *LHB* allele were largely healthy. The first report of this mutation among the male population was a patient with hypogonadotropic hypogonadism provided clinical and experimental evidence that this novel mutation causes selective LH deficiency.Other studies showed significantly higher serum LH concentrations in V-LH carriers among the young Baltic men and Estonian male infertility patients. This suggests polymorphism interaction with the genetic background.

Herein we described the presence of 5 single nucleotide polymorphisms of *LHB* gene, that shared between the investigated cases were with homozygous (AA) inheritance mode. They are located on the signal peptide and the homologue glycoprotein parts and thus could have significant influence on serum level of the hormone without inactivating or damaging its biological function. The most reported mutations of this research findings were (rs1800447 (W28R) and rs34349826 (I35T)) which are associated with protein glycosylation and have already been linked to response to controlled ovarian stimulation, male infertility and PCOS.

Protein glycosylation is one of the most frequent and relevant post-translational modifications and plays a key role in the modulation of protein properties particularly with regard to the half-life of the hormone, heterodimer stability and the intrinsic bioactivity. The carbohydrate content of LH is approximately 15%(N50) while that of FSH is 20%(N25,N42), and that of hCG is 30%(N33,N50) and the circulatory half-lives of the gonadotropins are about 20 mins for LH, 2 hours for FSH, and 12–24 hours for hCG. Molecular alterations of V-LHB, due to rs1800447 (Trp28Arg) and Ile35Thr (rs34349826) create an extra glycosylation signal (Asn-X-Thr) into the *LHB* chain, which introduces an oligosaccharide side chain into Asn13, similar to that present in HCG. This might increase the affinity binding between the peptide portion of the hormone and the receptor and also increase the lifespan of the hormone (Shafaghi*et al*., 2019).

Findings using serum variant homozygotes showed that V-LH is more active than WT-LH in bioassays in vitro, but it has a shorter half-life in circulation. The shorter half-life of V-LH is possibly compensated for by about (40%) higher promoter activity of the gene, explained by 8 SNPs in the (50-flanking regions) of *V-LHB*. They were reported as having a modulatory effect on the hyperandrogenemia of PCOS. More recently, Alviggi *et al*. have confirmed that *v-LHB* polymorphism produces a less active form of the hormone which is not able to support satisfactorily FSH activity during controlled ovarian stimulation, and that the *v-LH*β carriers experience a considerable decrease of the number of transferred embryos. The presence of this mutation statistically associated with high serum levels of LH and testosterone and irregular menstrual cycles highlighting the essential role of LH/FSH cooperation in the latest stages of follicle maturation.

The polymorphism rs5030775 (A18T) is associated with more potent stimulation of the IP3 response than wild type LH mapped on the signal peptide position, which is a short peptide located in the N-terminal of proteins which is responsible for protein recognition and secretion and it is often cleaved to generate the mature protein. Therefore, presence of this mutation within the leader sequence, which has been reported as potential stimulator of IP3 in contrast to the cAMP pathway, could cause an alteration of GnRH pulse frequency due to overstimulation of IP3. This in turn would increase *LHB* gene expression and might influence abnormal recognition of the prohormone within the endoplasmic reticulum when binding with SRP occurs. This would affect its interaction with the LHCGR receptor. Interestingly, a point mutation associated with autosomal recessive familial isolated hypoparathyroidism caused improper cleavage by signal peptidase at the normal position. Remarkably, this mutation ((historically reported as Thr-3A) A18T) was also found in 3% of the Rwanda population with a heterogeneous pattern that might be a sign of the evolutionarily background origin of PCOS and support the idea that a high level of serum *LHB* isassociated with increased pulse frequencies because of high GnRH pulse frequencies that favor *LHB* gene expression, whereas low GnRH pulse frequencies favor *FSHB* gene expression.

We are not aware of any literature in regarding to rs746167425 (R22K) and rs35270001 (H30R). With regards to their physiochemical properties Arginine mutated into a Lysine at position 22, which is smaller, would not be expected to affect protein stability. However, Histidine, located very close to a residue that makes a cysteine bond, forms a hydrogen bond with Tryptophan on position 28. When mutated to Arginine there is positive charge and a reduction in size. This creates an extra glycosylation site on the mutated formthat might cause loss of hydrophobic interactions with other molecules on the surface of the protein.

**In conclusion**, The diagnosis of PCOS makes the patient aware of possible fertility concerns, dysfunctional bleeding, endometrial cancer, obesity, diabetes, dyslipidemia, hypertension, and theoretical increased risk of cardiovascular disease. Since PCOS has genetic elements, knowledge of polymorphisms that affect LH action may bring awareness of PCOS to family members and future children. Given that insulin resistance is heavily associated with PCOS, with early genetic identification these individuals could be directed to increased screening to help reduce adverse diabetic and cardiovascular outcomes that will likely have better long-term outcomes. Interventions may include early lifestyle advice, as well as insulin sensitizing medication, such as metformin, screening for diabetes and hyperlipidemia. Overall, in Sudan there seem to be genetic markers that highlight the risk of PCOS that can be detected before PCOS is manifest.

## Data Availability

I Nidal Essa under signed, declare and affirm that this thesis is our own original work. I have followed all ethical and technical principles in the preparation, data collection, data analysis and compilation of this Thesis. Any scholarly matter that is included in the Thesis has been given recognition through citation. I, solemnly declare that this Thesis has not been submitted to any other institution anywhere for the award of any academic degree, diploma or certificate.

---

**Figure (1):**
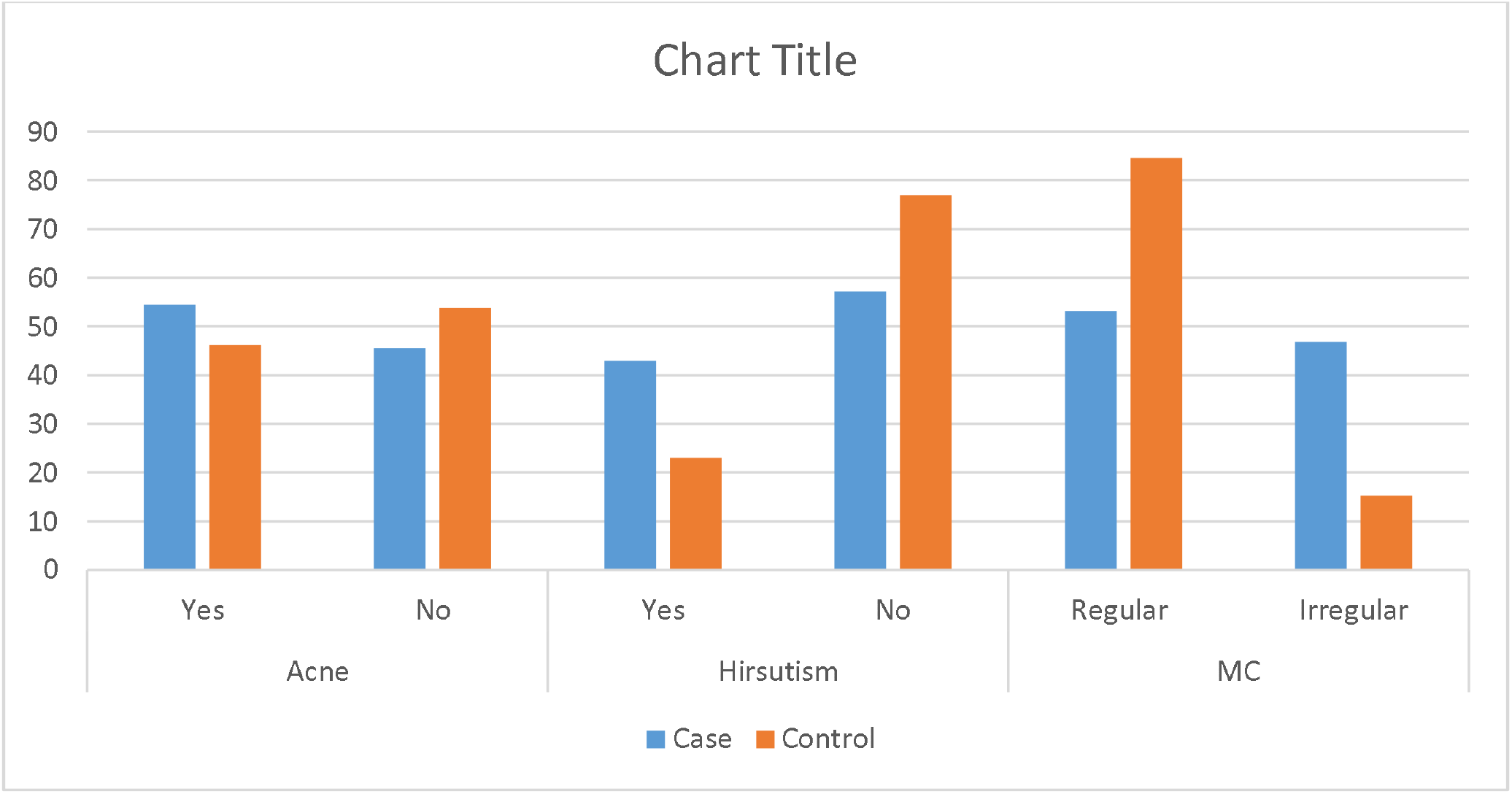
The frequencies of presence of Acne, hirsutism, and status of MC between study groups; (54, 5) but 57.1% of them had no facial hirsutism similar to 77% of control group.84.6% of our control had regular menstrual cycle and only 15.3 had irregular menstrual cycle, while the case group had the higher distribution than control group.

**Figure (2):**
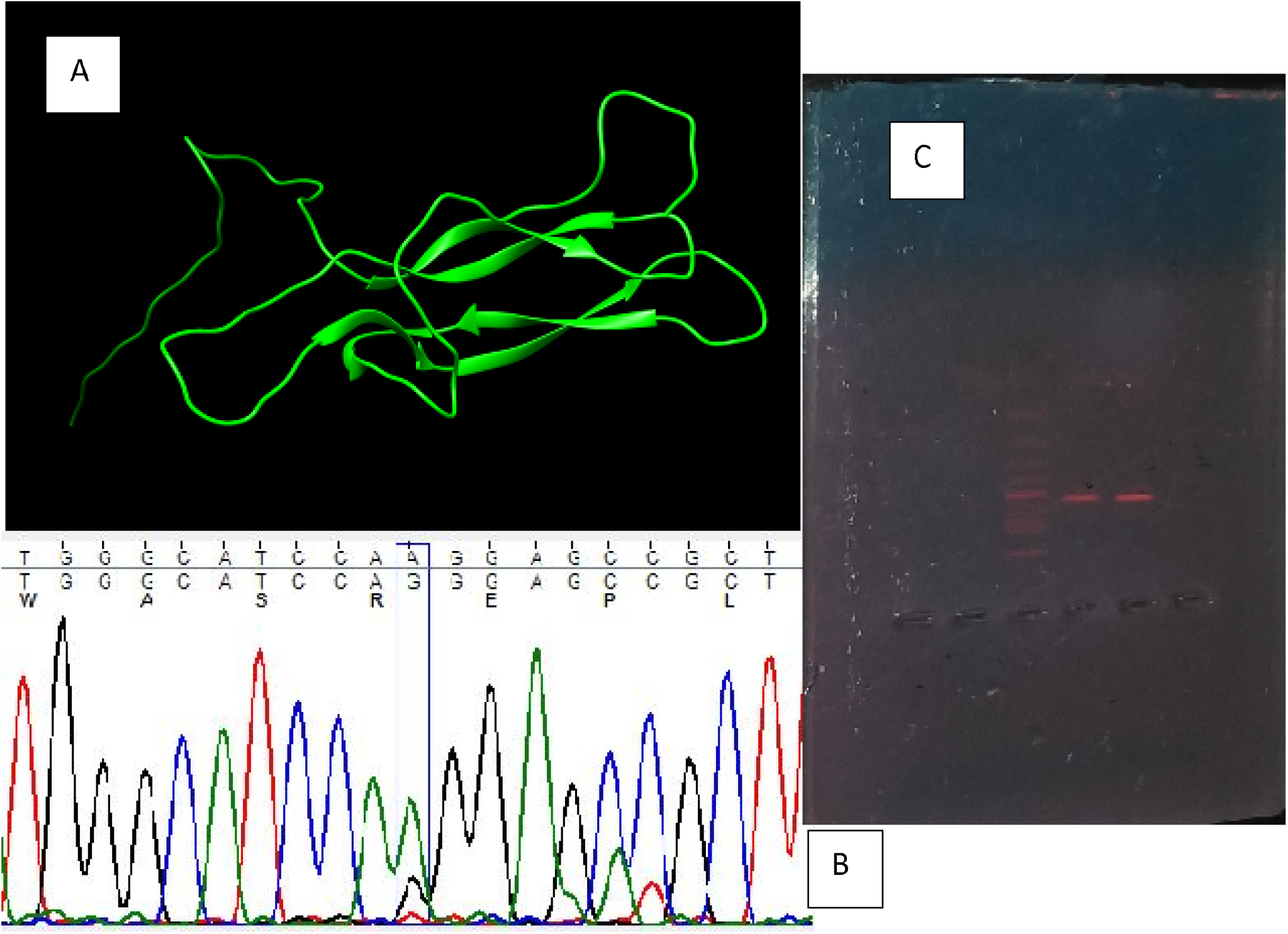
(A)The normal chemical structure of LH protein, (B) The chromatogram graph for the sequenced protein, (C) The gel electrophoresis showing the PCR product band (571bp).

**Figure (3):**
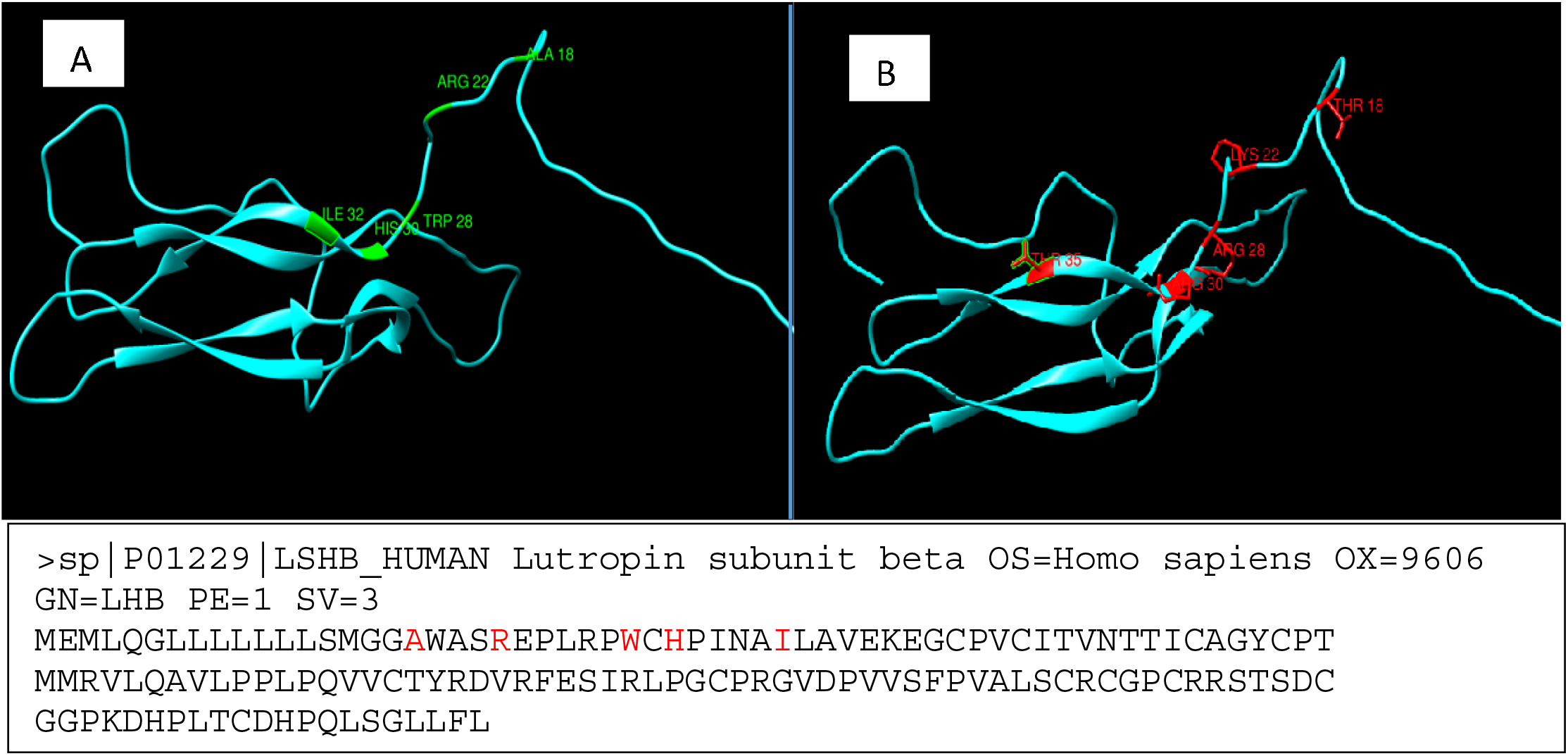
illustrated both normal and mutated amino acids on sequenced exon (2) LHβ gene

