## Supplemental table 1 for "Novel Mutations in Human Luteinizing Hormone Beta Subunit Related to Polycystic Ovary Syndrome among Sudanese Women"

>SeqK5 [organism=Homo sapiens] [isolate=EKS5] Homo sapiens, luteinizing hormone subunit beta (LHB), partial CDS BankIt2311635 5          MT041247

GGATCTGAAATGTTGGGGTATCTCAGGTCCTCTGGGCTGTGGGGTGGGCTCTGAAAGGCAGGTGTCCGGGTGGTGGGTCCTGAATAGGAGATGCCGGGAAGGGTCTCTGGGTCTTTGTGGGTGGTGTACCACGCGGGATGGGAAGGCCAGGACTCGGGGCTGCGGTCTCAGACCCGGGTGAAGCAGTGTCCTTGTCCCAGGGGCTGCTGCTGTTGCTGCTGCTGAGCATGGGCGGGACATGGGCATCCAAGGAGCCGCTTCGGCCACGGTGCCGCCCCATCAATGCCACCCTGGCTGTGGAGAAGGAGGGCTGCCCCGTGTGCATCACCGTCAACACCACCATCTGTGCCGGCTACTGCCCCACCATGGTGAGCTGCCCGGGGCCGGGGCAGGTGCTGCCACCTCAGGGCCAGACCCACAGAGGCAGCGGGGGAGGAAGGGTGGTCTGCCTCTCTGGTCAGGGGCTGCGGAATGGGGTGTGGGAGGGC

>SeqA6 [organism=Homo sapiens] [isolate= ALL6] Homo sapiens, luteinizing hormone subunit beta (LHB), partial CDS BankIt2311635 Seq        MT041245

GGATCTGAAATGTTGGGGTATCTCAGGTCCTCTGGGCTGTGGGGTGGGCTCTGAAAGGCAGGTGTCCGGGTGGTGGGTCCTGAATAGGAGATGCCGGGAAGGGTCTCTGGGTCTTTGTGGGTGGTGTACCACGCGGGATGGGAAGGCCAGGACTCGGGGCTGCGGTCTCAGACCCGGGTGAAGCAGTGTCCTTGTCCCAGGGGCTGCTGCTGTTGCTGCTGCTGAGCATGGGCGGGACATGGGCATCCAAGGAGCCGCTTCGGCCACGGTGCCGCCCCATCAATGCCACCCTGGCTGTGGAGAAGGAGGGCTGCCCCGTGTGCATCACCGTCAACACCACCATCTGTGCCGGCTACTGCCCCACCATGGTGAGCTGCCCGGGGCCGGGGCAGGTGCTGCCACCTCAGGGCCAGACCCACAGAGGCAGCGGGGGAGGAAGGGTGGTCTGCCTCTCTGGTCAGGGGCTGCGGAATGGGGTGTGGGAGGGC

>SeqN4 [organism=Homo sapiens] [isolate= NOO4] Homo sapiens, luteinizing hormone subunit beta (LHB), partial CDS BankIt2311657 Seq4       MT041251

GGATCTGAAATGTTGGGGTATCTCAGGTCCTCTGGGCTGTGGGGTGGGCTCTGAAAGGCAGGTGTCCGGGTGGTGGGTCCTGAATAGGAGATGCCGGGAAGGGTCTCTGGGTCTTTGTGGGTGGTGTACCACGCGGGATGGGAAGGCCAGGACTCGGGGCTGCGGTCTCAGACCCGGGTGAAGCAGTGTCCTTGTCCCAGGGGCTGCTGCTGTTGCTGCTGCTGAGCATGGGCGGGACATGGGCATCCAAGGAGCCGCTTCGGCCACGGTGCCGCCCCATCAATGCCACCCTGGCTGTGGAGAAGGAGGGCTGCCCCGTGTGCATCACCGTCAACACCACCATCTGTGCCGGCTACTGCCCCACCATGGTGAGCTGCCCGGGGCCGGGGCAGGTGCTGCCACCTCAGGGCCAGACCCACAGAGGCAGCGGGGGAGGAAGGGTGGTCTGCCTCTCTGGTCAGGGGCTGCGGAATGGGGTGTGGGAGGGC

>SeqB8 [organism=Homo sapiens] [isolate= BAN8] Homo sapiens, luteinizing hormone subunit beta (LHB), partial CDS BankIt2311635 8          MT041248

GGATCTGAAATGTTGGGGTATCTCAGGTCCTCTGGGCTGTGGGGTGGGCTCTGAAAGGCAGGTGTCCGGGTGGTGGGTCCTGAATAGGAGATGCCGGGAAGGGTCTCTGGGTCTTTGTGGGTGGTGTACCACGCGGGATGGGAAGGCCAGGACTCGGGGCTGCGGTCTCAGACCCGGGTGAAGCAGTGTCCTTGTCCCAGGGGCTGCTGCTGTTGCTGCTGCTGAGCATGGGCGGGACATGGGCATCCAAGGAGCCGCTTCGGCCACGGTGCCGCCCCATCAATGCCACCCTGGCTGTGGAGAAGGAGGGCTGCCCCGTGTGCATCACCGTCAACACCACCATCTGTGCCGGCTACTGCCCCACCATGGTGAGCTGCCCGGGGCCGGGGCAGGTGCTGCCACCTCAGGGCCAGACCCACAGAGGCAGCGGGGGAGGAAGGGTGGTCTGCCTCTCTGGTCAGGGGCTGCGGAATGGGGTGTGGGAGGGC

>SeqT9 [organism=Homo sapiens] [isolate= TAG9] Homo sapiens, luteinizing hormone subunit beta (LHB), partial CDS BankIt2311657 Seq9       MT041253

GGATCTGAAATGTTGGGGTATCTCAGGTCCTCTGGGCTGTGGGGTGGGCTCTGAAAGGCAGGTGTCCGGGTGGTGGGTCCTGAATAGGAGATGCCGGGAAGGGTCTCTGGGTCTTTGTGGGTGGTGTACCACGCGGGATGGGAAGGCCAGGACTCGGGGCTGCGGTCTCAGACCCGGGTGAAGCAGTGTCCTTGTCCCAGGGGCTGCTGCTGTTGCTGCTGCTGAGCATGGGCGGGACATGGGCATCCAAGGAGCCGCTTCGGCCACGGTGCCGCCCCATCAATGCCACCCTGGCTGTGGAGAAGGAGGGCTGCCCCGTGTGCATCACCGTCAACACCACCATCTGTGCCGGCTACTGCCCCACCATGGTGAGCTGCCCGGGGCCGGGGCAGGTGCTGCCACCTCAGGGCCAGACCCACAGAGGCAGCGGGGGAGGAAGGGTGGTCTGCCTCTCTGGTCAGGGGCTGCGGAATGGGGTGTGGGAGGGC

>SeqZ11 [organism=Homo sapiens] [isolate= ZEN11] Homo sapiens, luteinizing hormone subunit beta (LHB), partial CDS BankIt2311635 11         MT041246

GGATCTGAAATGTTGGGGTATCTCAGGTCCTCTGGGCTGTGGGGTGGGCTCTGAAAGGCAGGTGTCCGGGTGGTGGGTCCTGAATAGGAGATGCCGGGAAGGGTCTCTGGGTCTTTGTGGGTGGTGTACCACGCGGGATGGGAAGGCCAGGACTCGGGGCTGCGGTCTCAGACCCGGGTGAAGCAGTGTCCTTGTCCCAGGGGCTGCTGCTGTTGCTGCTGCTGAGCATGGGCGGGACATGGGCATCCAAGGAGCCGCTTCGGCCACGGTGCCGCCCCATCAATGCCACCCTGGCTGTGGAGAAGGAGGGCTGCCCCGTGTGCATCACCGTCAACACCACCATCTGTGCCGGCTACTGCCCCACCATGGTGAGCTGCCCGGGGCCGGGGCAGGTGCTGCCACCTCAGGGCCAGACCCACAGAGGCAGCGGGGGAGGAAGGGTGGTCTGCCTCTCTGGTCAGGGGCTGCGGAATGGGGTGTGGGAGGGC

>SeqF1 [organism=Homo sapiens] [isolate= FAT1] Homo sapiens, luteinizing hormone subunit beta (LHB), partial CDS BankIt2311657 Seq1       MT041255

GGATCTGAAATGTTGGGGTATCTCAGGTCCTCTGGGCTGTGGGGTGGGCTCTGAAAGGCAGGTGTCCGGGTGGTGGGTCCTGAATAGGAGATGCCGGGAAGGGTCTCTGGGTCTTTGTGGGTGGTGTACCACGCGGGATGGGAAGGCCAGGACTCGGGGCTGCGGTCTCAGACCCGGGTGAAGCAGTGTCCTTGTCCCAGGGGCTGCTGCTGTTGCTGCTGCTGAGCATGGGCGGGACATGGGCATCCAAGGAGCCGCTTCGGCCACGGTGCCGCCCCATCAATGCCACCCTGGCTGTGGAGAAGGAGGGCTGCCCCGTGTGCATCACCGTCAACACCACCATCTGTGCCGGCTACTGCCCCACCATGGTGAGCTGCCCGGGGCCGGGGCAGGTGCTGCCACCTCAGGGCCAGACCCACAGAGGCAGCGGGGGAGGAAGGGTGGTCTGCCTCTCTGGTCAGGGGCTGCGGAATGGGGTGTGGGAGGGC

>SeqSOLHB [organism=Homo sapiens] [isolate= SOM3] Homo sapiens, luteinizing hormone subunit beta (LHB), partial CDS BankIt2311635 3          MT041244

GGCTGCGGTCTCAGACCCGGGTGAAGCAGTGTCCTTGTCCCAGGGGCTGCTGCTGTTGCTGCTGCTGAGCATGGGCGGGACATGGGCATCCAAGGAGCCGCTTCGGCCACGGTGCCGCCCCATCAATGCCACCCTGGCTGTGGAGAAGGAGGGCTGCCCCGTGTGCATCACCGTCAACACCACCATCTGTGCCGGCTACTGCCCCACCATGGTGAGCTGCCCGGGGCCGGGGCAGGTGCTGCCACCTCAGGGCCAGACCCACAGAGGCAGCGGGGGAGGAAGGGTGGTCTGCCTCTCTGGTCAGGGGCTGCGGAATGGGGTGTGGGAGGGC
